# Cost-effectiveness of vitamin D_3_ supplementation in older adults with vitamin D deficiency in Ireland

**DOI:** 10.1101/2021.10.31.21265715

**Authors:** Laurence F. Lacey, David J. Armstrong, Emily Royle, Pamela J. Magee, L. Kirsty Pourshahidi, Sumantra Ray, J. J. Strain, Emeir M. McSorley

**Affiliations:** Lacey Solutions Ltd, Skerries, Co. Dublin, Ireland; Nutrition Innovation Centre for Food and Health (NICHE), Ulster University, Coleraine, BT52 1SA, United Kingdom; Department of Rheumatology, Western Health and Social Care Trust, Londonderry, UK; NNEdPro Global Centre for Nutrition and Health in Cambridge, UK

**Keywords:** vitamin D deficiency, vitamin D_3_ supplementation, older adults, public health programme, primary prevention, mortality, fractures, health economics, cost-effectiveness

## Abstract

**Background:** This study investigated the cost-effectiveness of vitamin D_3_ supplementation in older adults in Ireland, with year-round vitamin D deficiency (serum 25-hydroxyvitamin D concentration <30 nmol/L) (13% of Irish adults), from the perspective of the Health Service Executive (HSE).

**Methods:** Three age groups were investigated: (1) ≥50 years, (2) ≥60 years, (3) ≥70 years. Based on the clinical literature, vitamin D_3_ supplementation may (1) decrease all-cause mortality by 7%, and (2) reduce hip fractures by 16%, and non-hip fractures by 20%. A discount rate of 4% was applied to life years and QALYs gained. The annual healthcare costs per patient used in the model are based on the average annual health resource use over the 5-year time horizon of the model.

**Results:** The cost/QALY estimates in all three age groups are below the usually acceptable cost-effectiveness threshold of €20,000/QALY. The most cost-effective and least costly intervention was in adults ≥70 years. For this age group, the average annual costs and outcomes would be approx. €6.2 million, 1,043 QALYs gained, with a cost/QALY of approx. €6,000. The results are most sensitive to the mortality risk reduction following vitamin D_3_ supplementation.

**Conclusion:** The cost-effectiveness of vitamin D_3_ supplementation is most robust in adults ≥70 years. Clinical uncertainty in the magnitude of the benefits of vitamin D_3_ supplementation could be further addressed by means of (1) performing a clinical research study or (2) conducting a pilot/regional study, prior to reaching a decision to invest in a full nationwide programme.

## INTRODUCTION

Clinical vitamin D deficiency (serum 25-hydroxyvitamin D (25(OH)D) concentration below <30 nmol/L) increases the risk of excess mortality and disease [1]. Vitamin D_3_ supplementation is likely to be clinically most beneficial in deficiency [1]. Vitamin D deficiency, as measured by serum 25(OH)D, is particularly high amongst older Irish adults [2].

There is a growing interest in the cost-effectiveness of vitamin D to prevent disease. While several previous studies were mainly conducted in the elderly for fall and/or fractures (e.g., [3,4]), more recently, a study estimated the costs and savings for preventing cancer deaths by vitamin D supplementation of the population aged ≥50 years in Germany [5]. The results of this study supported the use of vitamin D_3_ supplementation among older adults as a potential cost saving approach to substantially reduce cancer mortality [5].

The objective of this study is to investigate the cost-effectiveness of vitamin D_3_ supplementation in older adults in Ireland, with year-round vitamin D deficiency (25(OH)D concentration <30 nmol/L) (13% of adults [2]), from the perspective of the Health Service Executive (HSE).

## METHODS

### Deaths and fractures avoided

The methodology used builds upon that used by Niedermaier, et al. [5]. The methodology applied to Ireland uses Central Statistics Office (CSO) Irish Life Table No. 17, for males and females, for the period, 2015-2017 [6]. The data for male and females from the Life Table were combined using Irish population statistics for 2016 [7] and applying the method of weighted averaging. The annual mortality probability and the expected life expectancy were determined for all ages. A 4% annual discount rate [8] was applied in order to calculate the discounted life expectancy. The “at risk” population of interest (i.e., those with year-round vitamin D deficiency) is estimated to be 13% of the Irish population [2]. This population cohort was estimated by applying 13% to the Irish population statistics for 2016 for all ages. The population data were summarised into 5-year age groups, beginning at age 50 years. The final age group was for those ≥85 years (with a mean age assumed to be 90 years, when required). For any given 5-year age group, the expected annual number of deaths is equal to the number of people in the age group multiplied by the annual mortality probability for the mid-point age of the group.

Based on the clinical literature, vitamin D_3_ supplementation may (1) decrease all-cause mortality by 7% [9], of which 4.2% (of the 7%) is a reduction in cancer mortality, and (2) reduce hip fractures by 16% (and related excess mortality) [10], and non-hip fractures by approx. 20% [11,12]. In the base-case analysis, the reduction in all-cause mortality was implemented in the model by applying a 7% reduction to the expected number of deaths occurring in each 5-year age group. This point-estimate reduction in all-cause mortality was varied over the range 1% to 10% in the univariate sensitivity analysis.

The number of hip fractures in 2019 and their distribution by age were obtained from the Hip Fracture database [13]. The number of non-hip fractures in 2014 was estimated from the total number of fractures and the number of hip fractures reported in the literature [14]. The number of non-hip fractures was assumed to have the same relative age distribution, as was found for hip fractures. It was conservatively assumed that only 13% of people who have a fracture would have year-round vitamin D deficiency and be included in the “at risk” population of interest [2]. The reductions in hip fractures (16%, and the associated estimated 1-year mortality of 22% following a hip fracture [15]) and non-hip fractures (approx. 20%) were included in the base-case analysis and varied in the univariate sensitivity analysis.

The results obtained from the model were investigated for three age cohorts: (1) ≥50 years, (2) ≥60 years, (3) ≥70 years.

### Discounted life years and discounted QALYs gained

The number of discounted life years gained for any given age group is equal to the number of deaths avoided multiplied by the associated discounted life expectancy. The number of discounted QALYs gained for any given age group is equal to number of discounted life years gained multiplied by the age-related utility value for the mid-point of the age group.

EQ-5D index population norms for the UK [16] were used for age-related utility values. The following age-related utility equation was used, for ages ≥50 years (which had a coefficient of determination (R^2^) of approx. 99%).

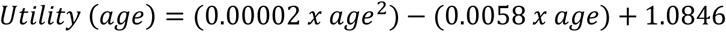

The disutility estimate used for hip-fracture was 0.20 and for non-hip-fracture was 0.09 [4]. These disutility estimates were applied for one year to estimate fracture related QALYs gained, i.e., number of fractures avoided per annum multiplied by the relevant fracture disutility.

### Comparator and time horizon

The comparator is the current standard of care, which is being compared against a systematic public health programme (1) to identify year-round vitamin D deficient adults (≥50 years), by means of serum 25(OH)D measurement, and (2) to treat such adults with vitamin D_3_ supplementation, under GP supervision, to render the patients vitamin D replete and to maintain their vitamin D adequacy over time.

A 5-year time horizon was employed in the sense that it was assumed that it would take 5-years for the public health programme to have identified the eligible vitamin D deficient patients and for them to have been treated for their vitamin D deficiency, and to be maintained in a vitamin D replete state.

### Healthcare resource use and costs

For each age-group, it was assumed that the average annual healthcare cost would be the cost of treating all the eligible patients plus the cost of patient identification (i.e., a blood 25(OH) vitamin D measurement in order to identify the “at risk” population), with the latter “set-up” cost averaged over 5-years. Thus, only those patients who are GP-assessed to be year-round vitamin D deficient are subsequently treated with vitamin D_3_ supplementation and monitored. Vitamin D deficiency was assumed to be treated with vitamin D_3_ 4,000 IU daily for up to 10 weeks, followed by 800 IU daily thereafter. Healthcare costs included in the analysis are: serum 25(OH)D measurement, the average cost of a GP visit, and drug acquisition costs of vitamin D_3_ supplements.

The cost off-sets included in the analysis were end-of-life invasive cancer costs avoided and the costs of fractures (hip and non-hip) avoided. The reduction in all-cause mortality was implemented in the model by applying a 7% reduction to the expected number of deaths occurring in each 5-year age group. This 7% reduction in all-cause mortality could be “partitioned” into a 4.2% reduction in cancer mortality and a 2.8% reduction in non-cancer mortality [9]. Thus, approx. 61% of the reduction in mortality was due to the reduction in cancer mortality. Invasive cancers constitute approx. 55% of all cancers [17]. Thus, approx. 33% of the reduction in mortality was due to the reduction in invasive cancer mortality. Conservatively, for deaths avoided, end-of-life costs of invasive cancers avoided were the only cost off-sets included in the analysis.

Average UK end-of-life cancer costs [18] were applied to Irish invasive cancer deaths, using the method of purchasing power parity (PPP), with inflation of healthcare costs to €(2020) values. Additional healthcare cost off-sets included in the analysis were due to the healthcare costs of hip and non-hip fractures avoided. The unit costs of all healthcare resource use items included in the model are summarised in Table 1.

**Table 1:**
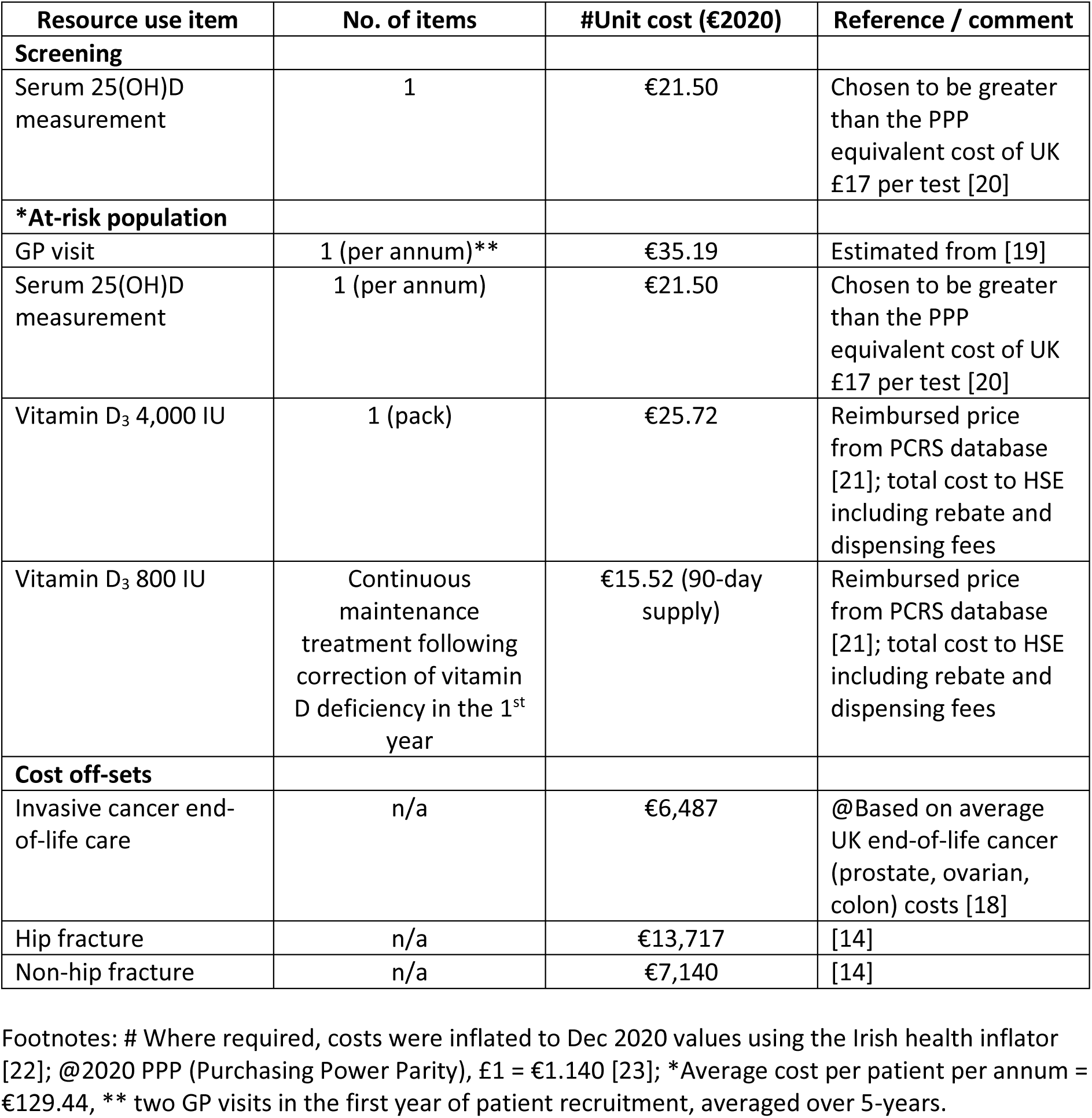
Healthcare resource use items and associated unit costs.

| Resource use item | No. of items | #Unit cost (€2020) | Reference / comment |
| --- | --- | --- | --- |
| **Screening** |  |  |  |
| Serum 25(OH)D measurement | 1 | €21.50 | Chosen to be greater than the PPP equivalent cost of UK £17 per test [20] |
| **\*At-risk population** |  |  |  |
| GP visit | 1 (per annum)** | €35.19 | Estimated from [19] |
| Serum 25(OH)D measurement | 1 (per annum) | €21.50 | Chosen to be greater than the PPP equivalent cost of UK £17 per test [20] |
| Vitamin D<sub>3</sub> 4,000 IU | 1 (pack) | €25.72 | Reimbursed price from PCRS database [21]; total cost to HSE including rebate and dispensing fees |
| Vitamin D<sub>3</sub> 800 IU | Continuous maintenance treatment following correction of vitamin D deficiency in the 1<sup>st</sup> year | €15.52 (90-day supply) | Reimbursed price from PCRS database [21]; total cost to HSE including rebate and dispensing fees |
| **Cost off-sets** |  |  |  |
| Invasive cancer end-of-life care | n/a | €6,487 | @Based on average UK end-of-life cancer (prostate, ovarian, colon) costs [18] |
| Hip fracture | n/a | €13,717 | [14] |
| Non-hip fracture | n/a | €7,140 | [14] |

Footnotes: # Where required, costs were inflated to Dec 2020 values using the Irish health inflator [22]; @2020 PPP (Purchasing Power Parity), £1 = €1.140 [23]; \*Average cost per patient per annum = €129.44, \*\* two GP visits in the first year of patient recruitment, averaged over 5-years.

The cost-effectiveness analysis did not include any discounting of healthcare costs.

### Univariate and multi-variate sensitivity analyses

The univariate sensitivity analyses and multi-variate sensitivity analysis performed are summarised in Table 2.

**Table 2:** Parameterisation of the univariate and multi-variate sensitivity analyses undertaken.

| **Parameter for univariate sensitivity analyses** | **Basecase estimate** | **Range or measure of variation evaluated** |
| --- | --- | --- |
| % Reduction in all-cause mortality | 7% | 1% to 10% |
| The risk reduction of fractures (hip and non-hip) | 16% (hip)<br>20% (non-hip) | 1% to 150% (of the basecase estimates) |
| Discount rate | 4% | 0% to 6% |
| Serum 25(OH)D measurement cost | €21.50 | 50% increase in the basecase estimate |
| Invasive cancer end-of-life care cost | €6,487 | 50% increase in the basecase estimate |
| Average annual cost of serum 25(OH)D measurement, GP visit, plus vitamin D acquisition cost | €129.44 | 50% increase in the basecase estimate |
| **Parameterisation of multi-variate sensitivity analysis** | **Basecase estimate** | **Estimates used in multi-variate sensitivity analysis** |
| % Reduction in all-cause mortality | 7% | 4.9%<br>(30% reduction from basecase) |
| The risk reduction of fractures (hip and non-hip) | 16% (hip)<br>20% (non-hip) | 11% (hip)<br>14% (non-hip)<br>(30% reduction from basecase) |
| Serum 25(OH)D measurement cost | €21.50 | 30% increase in the basecase estimate |
| Invasive cancer end-of-life care cost | €6,487 | 30% increase in the basecase estimate |
| Average annual cost of serum 25(OH)D measurement, GP visit, plus vitamin D acquisition cost | €129.44 | 30% increase in the basecase estimate |

All results of the data analysis given below were obtained using Microsoft Excel 2019, 32-bit version.

## RESULTS

### Basecase results

The basecase cost-effectiveness results are summarised in Table 3. The cost/QALY estimates in all three age groups are below the usually acceptable cost-effectiveness threshold of €20,000/QALY in Ireland. The most cost-effective and least costly intervention was in adults ≥70 years of age. While a public health primary prevention programme in adults ≥70 years of age would produce fewer QALYs, it would be sufficiently less costly to result in the lowest (best) cost/QALY.

**Table 3:** Basecase cost-effectiveness of vitamin D_3_ supplementation in older adults with vitamin D deficiency plus results of uni-variate and multi-variate sensitivity analyses.

| Description | Age Group | Net average cost of vit D deficiency treatment programme per annum | Discounted QALYs gained | Cost-effectiveness (Cost/QALY) |
| --- | --- | --- | --- | --- |
| Basecase | ≥ 50 years | €27,334,061 | 1,690 | €16,177 |
|  | ≥ 60 years | €15,327,744 | 1,446 | €10,599 |
|  | ≥ 70 years | €6,233,810 | 1,043 | €5,976 |
| increasing the serum 25(OH)D measurement cost by 50% |  |  |  |  |
|  | ≥ 50 years | €32,465,378 | 1,690 | €19,214 |
|  | ≥ 60 years | €18,436,854 | 1,446 | €12,749 |
|  | ≥ 70 years | €7,746,219 | 1,043 | €7,426 |
| increasing the average end-of-life invasive cancer cost by 50% |  |  |  |  |
|  | ≥ 50 years | €27,051,323 | 1,690 | €16,010 |
|  | ≥ 60 years | €15,063,639 | 1,446 | €10,417 |
|  | ≥ 70 years | €6,008,899 | 1,043 | €5,760 |
| increasing the average annual cost of serum 25(OH)D measurement, GP visit, plus vitamin D<sub>3</sub> supplementation acquisition cost by 50% |  |  |  |  |
|  | ≥ 50 years | €39,504,375 | 1,690 | €23,380 |
|  | ≥ 60 years | €22,701,845 | 1,446 | €15,699 |
|  | ≥ 70 years | €9,820,899 | 1,043 | €9,414 |
| “pessimistic” multi-variate sensitivity analysis |  |  |  |  |
|  | ≥ 50 years | €38,930,185 | 1,183 | €32,907 |
|  | ≥ 60 years | €22,686,070 | 1,013 | €22,405 |
|  | ≥ 70 years | €10,143,360 | 729 | €13,918 |

Abbreviations: vit D, vitamin D; QALY, quality adjusted life year; 25(OH)D, serum 25-hydroxyvitamin D

All additional basecase results will be focused on those obtained in adults ≥70 years of age.

The annual healthcare cost offsets reduce the average annual costs from a total of approximately €9.0 million to approximately €6.2 million per annum for treating older adults ≥ 70 years, who are year-round vitamin D deficient, with vitamin D_3_ supplementation (Figure 1).

**Figure 1:**
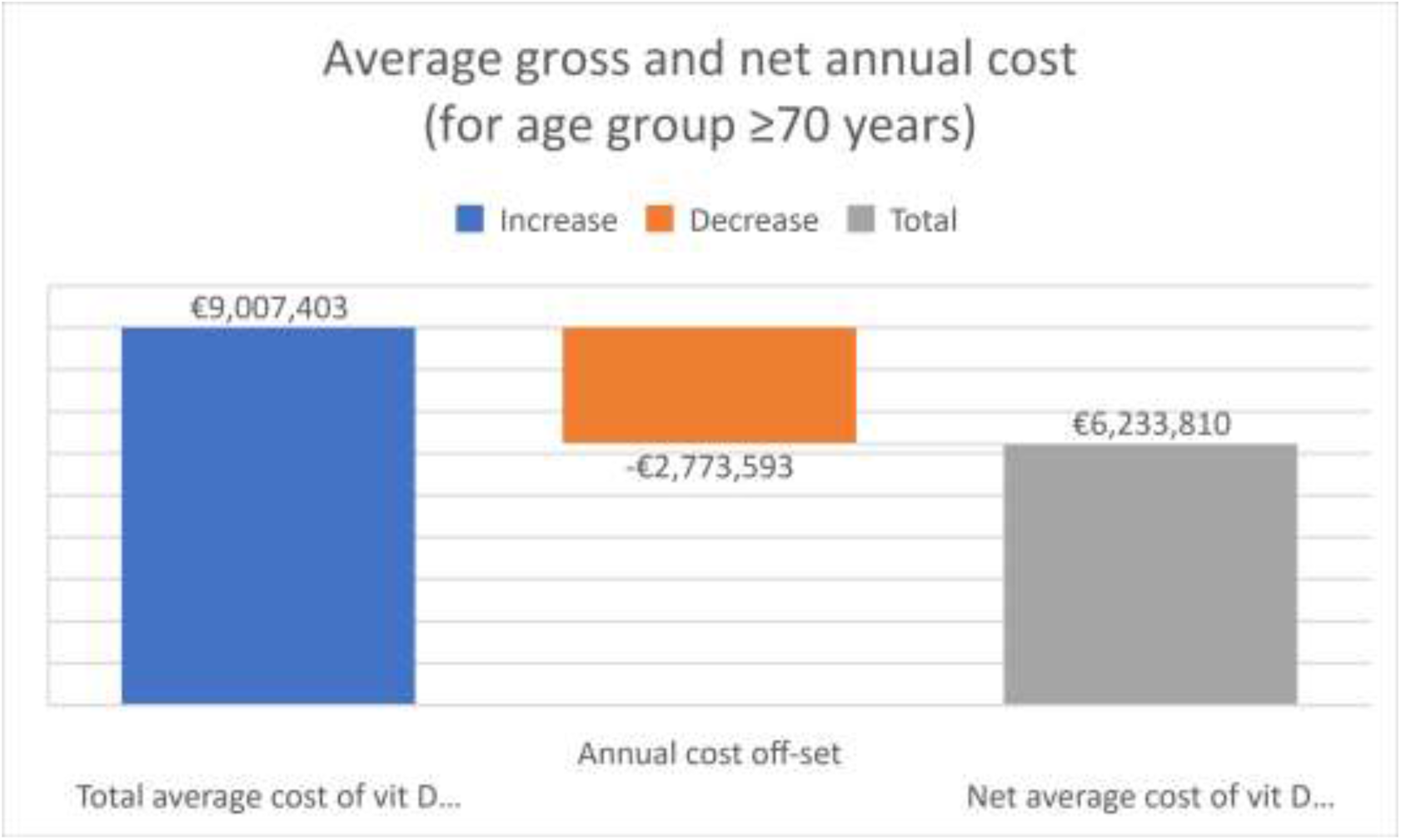
Waterfall plot of basecase average annual total, cost off-set, and net healthcare costs in older adults ≥ 70 years. Abbreviations: vit D, vitamin D

Vitamin D_3_ supplementation drug acquisition costs are the biggest cost component, being approximately 41% of the total €9.0 million annual healthcare costs (excluding cost offsets) (Figure 2). Approximately 84% of the annual healthcare cost off-sets are the healthcare costs of fractures avoided, with the remaining approximate 16% of the annual healthcare cost off-sets due to end-of-life care costs avoided from a reduction in invasive cancer deaths.

**Figure 2:**
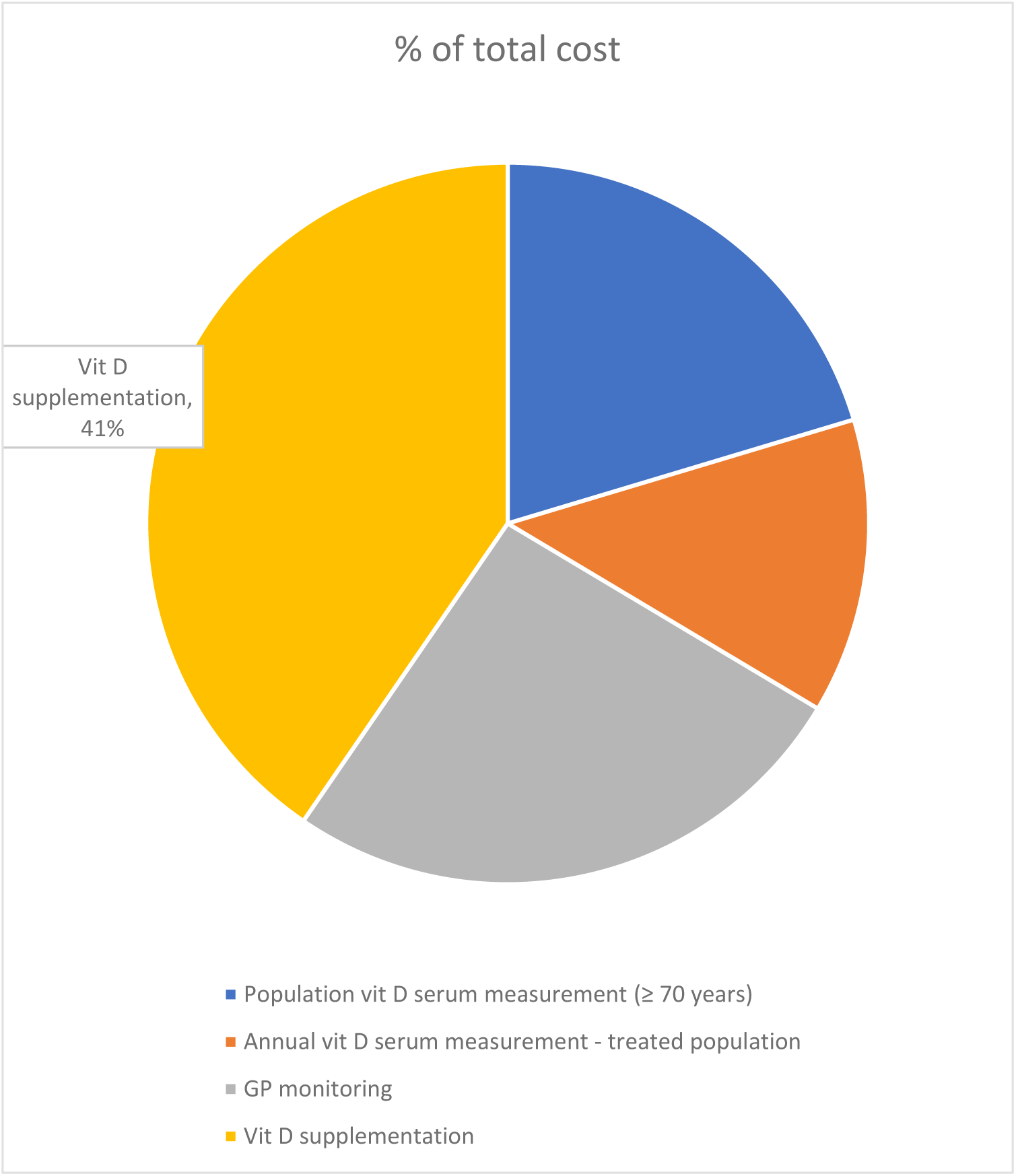
Composition of the basecase average annual total annual costs (excluding cost off-sets) in older adults ≥ 70 years. Abbreviations: vit D, vitamin D

### Results of the sensitivity analyses

The impact on the cost/QALY of varying the % reduction in all-cause mortality from treating older adults, who are year-round vitamin D deficient, with vitamin D_3_ supplementation is given in Figure 3. As can be seen, for elderly adults ≥70 years of age, even if the reduction in all-cause mortality due to Vitamin D_3_ supplementation was reduced to 2% (7% in the basecase), the cost/QALY in this age group would still be less than €20,000 per QALY gained. A similar threshold analysis can be performed for the other age-groups by visible inspection of Figure 3.

**Figure 3:**
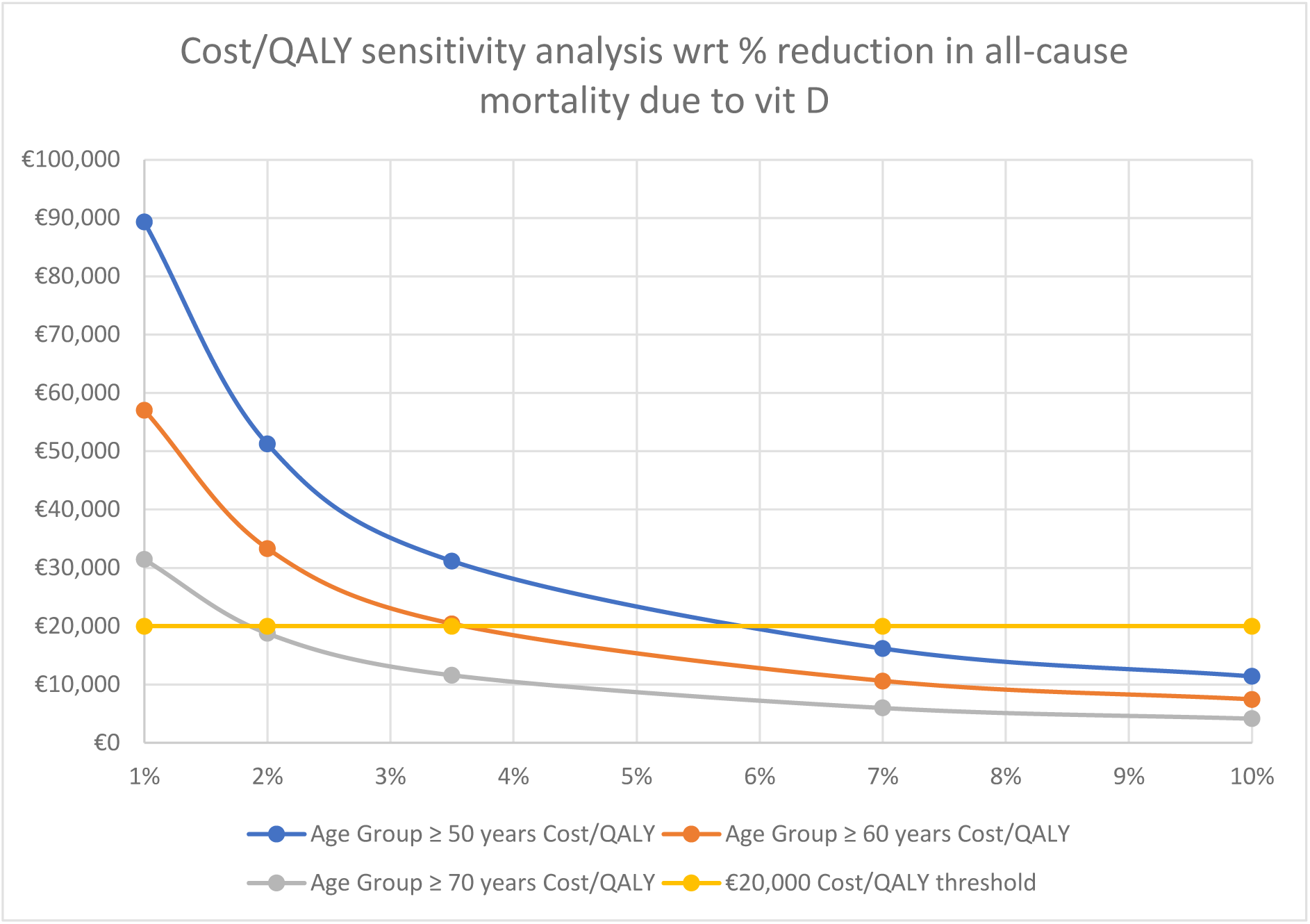
Univariate sensitivity analysis: impact on the cost/QALY of varying the % reduction in all-cause mortality from treating those older adults, who are year-round vitamin D deficient, with vitamin D_3_ supplementation (basecase = 7% reduction in all-cause mortality) Abbreviations: wrt, with respect to; vit D, vitamin D; QALY, quality adjusted life year

The impact on the cost/QALY of varying the risk reduction of fractures (hip and non-hip) from treating older adults, who are year-round vitamin D deficient, with vitamin D_3_ supplementation was investigated; in all cases explored, the cost/QALY remain less than €20,000 per QALY gained, for each of the three age-groups (Figure 4).

**Figure 4:**
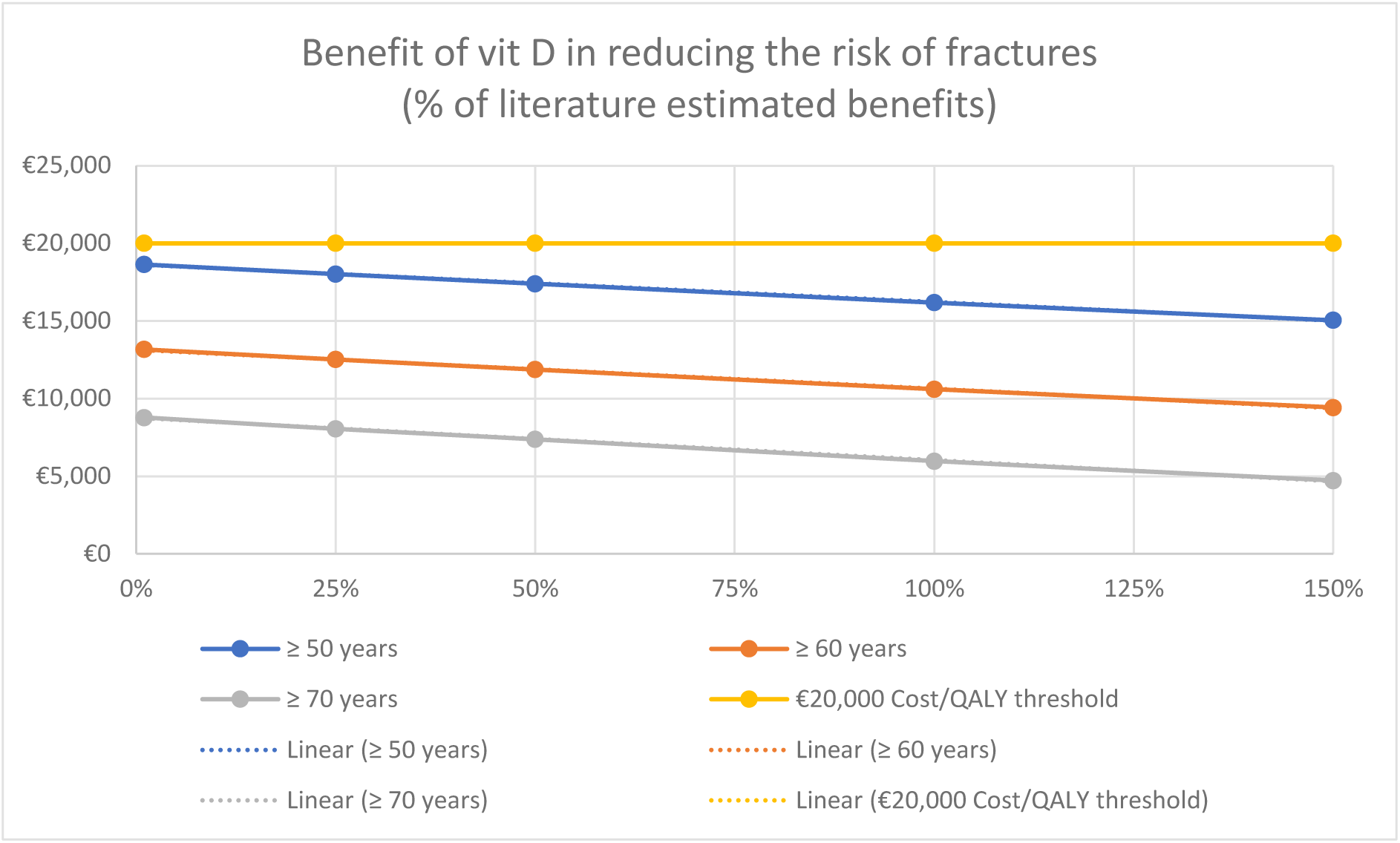
Univariate sensitivity analysis: impact on the cost/QALY of varying the risk reduction of fractures (hip and non-hip) from the published literature as a result of treating older adults, who are year-round vitamin D deficient, with vitamin D_3_ supplementation (i.e., the risk reduction estimates were varied over the range 1% to 150% of the literature estimates used in the basecase) Abbreviations: vit D, vitamin D; QALY, quality adjusted life year

The impact on the cost/QALY of varying the discount rate per annum (4% in the basecase) was investigated; in all cases explored, the cost/QALY remain less than €20,000 per QALY gained, for all age-groups (Figure 5).

**Figure 5:**
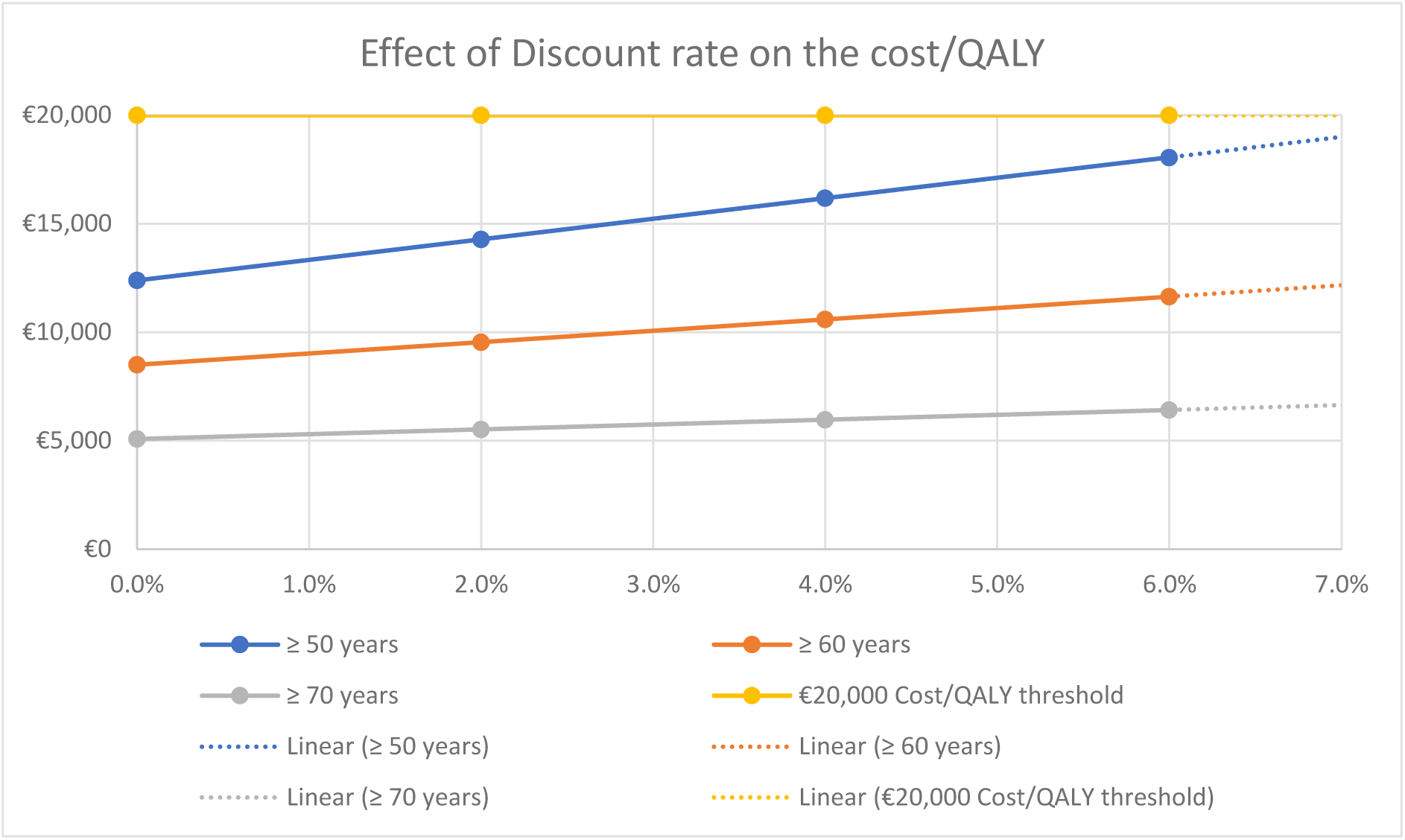
Univariate sensitivity analysis: impact on the cost/QALY of varying the discount rate for treating older adults, who are year-round vitamin D deficient, with vitamin D_3_ supplementation (basecase = 4% discount rate per annum) Abbreviations: vit D, vitamin D; QALY, quality adjusted life year

In terms of healthcare costs, the most uncertain are in relation to (1) serum 25(OH)D measurement, and (2) average end-of-life invasive cancer cost. In a univariate sensitivity analysis:

1. The serum 25(OH)D measurement cost was increased by 50% (compared to the basecase, from €21.50 to €32.25), which will tend to increase the cost per QALY results (Table 3). While the cost per QALY results increased modestly for each age cohort compared to the basecase results, but remain below the €20,000/QALY threshold (Table 3).
2. The average end-of-life invasive cancer cost was increased by 50% (compared to the basecase, from €6,487 to €9,731), as it was based on a 2005 reference [17]. This cost off-set will tend to decrease the cost per QALY results (Table 3).

The cost per QALY results were not very sensitive to large changes (50% increase) in either of these uncertain unit costs.

Potentially, some vitamin D deficient elderly adults may require additional serum 25(OH)D measurement, GP visit(s) per annum, and/or additional course(s) of vitamin D_3_ supplementation to correct their vitamin D deficiency. To investigate this, a sensitivity analysis was conducted in which the average annual cost of serum 25(OH)D measurement, GP visit, plus vitamin D_3_ supplementation acquisition cost (€129.44 in the basecase) was increased by 50% (to €194.17) (Table 3). In this case, the cost per QALY results increased for each age cohort compared to the basecase results; however, for both the age groups: ≥60 years and ≥70 years, the cost per QALY were well below €20,000/QALY (Table 3).

### Results of a “pessimistic” scenario

The parameterisation for a multi-variate sensitivity analysis performed is summarised in Table 2. In this case, the potential clinical benefits of vitamin D_3_ supplementation to vitamin D deficient elderly adults are decreased by 30% compared to the basecase, whereas, the relevant healthcare costs are increased by 30%. This is regarded as a “pessimistic” scenario.

In this “pessimistic” scenario, the cost per QALY for the age group: ≥70 years remained well below €20,000/QALY, whereas, this was no longer the case for the age group: ≥60 years, with the cost per QALY considerably above €20,000/QALY for the age group: ≥50 years (Table 3).

## DISCUSSION

Vitamin D_3_ supplementation is likely to be clinically most beneficial in deficiency [1]. Furthermore, those most likely to gain maximum benefit from vitamin D_3_ supplementation are those, with year-round vitamin D deficiency (25(OH)D concentration <30 nmol/L), which is estimated to be 13% of Irish adults [2]. What has been evaluated in this paper is the healthcare costs and benefits of (1) identifying year-round vitamin deficient adults (≥50 years), by means of serum 25(OH)D measurement, and (2) treating such adults with vitamin D_3_ supplementation, under GP supervision, to render the patients vitamin D replete and to maintain their vitamin D adequacy over time. It is envisaged that GP supervision could be performed through routine GP / patient interaction. However, it was also assumed that those patients identified as having year-round vitamin D deficiency would require, on average, one additional GP visit per annum, with a further additional GP visit in the year that the qualifying patient would enter the potential public health programme. It is further assumed that serum 25(OH)D measurement during the summer the summer months is sufficient to identify those older adults with year-round vitamin D deficiency. No calcium supplements were included in the cost-effectiveness analysis, as it was assumed that adequate calcium intake by patients could be achieved through dietary advice provided as part of the GP visit.

The cost-effectiveness modelling assumed that patient identification and treatment with vitamin D_3_ supplementation would take some years to be achieved. Also, the benefits of treating vitamin D deficiency would take time to be manifested in terms of clinical benefits. Therefore, it was assumed that a “steady-state” would be achieved by the end of 5-years from initiation of the public health programme. For each age-group, it was assumed that the average annual healthcare cost would be the cost of treating all the eligible patients plus the cost of patient identification, with the latter “set-up” costs averaged over 5-years. It is unknown, however, whether this time frame would be sufficient to capture the potential benefits of vitamin D_3_ supplementation on reducing all-cause mortality or reducing the risk of fractures.

The cost/QALY estimates in all three age groups are below the usually acceptable cost-effectiveness threshold of €20,000/QALY in Ireland. The most cost-effective and least costly intervention was in adults ≥70 years of age. While a public health primary prevention programme in adults ≥70 years of age would produce fewer QALYs, it would be sufficiently less costly to result in the lowest (best) cost/QALY. Therefore, it is proposed that a GP-monitored, vitamin D_3_ supplementation public health programme be considered in adults ≥70 years of age, in the first instance.

The cost-effectiveness results could potentially be improved if additional clinical benefits of vitamin D_3_ supplementation had been included in the model, e.g., decreasing the incidence of respiratory tract infections and the adverse clinical consequences than can arise from such infections [24]. In addition, a conservative approach was taken where applicable. For example, the cost off-sets due to cancer end-of-life care were limited to invasive cancer care costs only, with invasive cancers constituting approx. 55% of all cancers [17]. In addition, discounting was applied to healthcare benefits (life years / QALYs gained from deaths avoided), but not to healthcare costs, such as the “set-up” costs, involving serum 25(OH)D measurement.

The results of the cost effectiveness analysis are most sensitive to the mortality risk reduction following vitamin D_3_ supplementation. For elderly adults ≥70 years of age, it was found that even if the reduction in all-cause mortality due to vitamin D_3_ supplementation was reduced to 2% (7% in the basecase), the cost/QALY in this age group would still be less than €20,000 per QALY gained. However, potentially the reduction in all-cause mortality due to vitamin D_3_ supplementation could be less than this.

Typically, the uncertainty in cost-effectiveness analysis can be assessed through probabilistic sensitivity analysis (PSA). A PSA was is not performed in this study. However, the nature of the clinical evidence base for vitamin D_3_ supplementation may not be very suitable for clinical uncertainty to be assessed in such a manner. The clinical uncertainty might be better addressed by means of (1) performing a clinical research study prior to reaching a decision to invest in a GP-monitored, vitamin D_3_ supplementation public health programme or (2) conducting a pilot/regional study prior to reaching a decision to invest in a full nationwide programme.

## CONCLUSION

The cost/QALY estimates of a GP-monitored, vitamin D_3_ supplementation public health programme in the three age groups: (1) ≥50 years, (2) ≥60 years, and (3) ≥70 years, are below the usually acceptable cost-effectiveness threshold of €20,000/QALY in Ireland. The cost-effectiveness of vitamin D_3_ supplementation is most robust in adults ≥70 years. The results of the cost effectiveness analysis are most sensitive to the mortality risk reduction following vitamin D_3_ supplementation. It is proposed that a GP-monitored, vitamin D_3_ supplementation public health programme be considered in adults ≥70 years of age, in the first instance.

## Data Availability

The methodology used is described in detail in the manuscript. The health economic analysis is available as a MS Excel spreadsheet

## Contributors

All authors reviewed and verified the final manuscript for submission. All authors were involved in the design and interpretation of the economic evaluation, although the analysis was carried out by LFL.

## Patient consent for publication

Not applicable.

## Ethics approval

Not applicable.

## REFERENCES

1. Amrein K, Scherkl M, Hoffmann M, et al. Vitamin D deficiency 2.0: an update on the current status worldwide (2020). Eur. J. Clin. Nutr; 74: 1498–1513. url https://www.nature.com/articles/s41430-020-0558-y

2. McCartney DM, Byrne DG. Optimisation of Vitamin D Status for Enhanced Immuno-protection Against Covid-19 (2020). Ir Med J; 113(4): P58. url https://imj.ie/wp-content/uploads/2020/04/Optimisation-of-Vitamin-D-Status-for-Enhanced-Immuno-protection-Against-Covid-19.pdf

3. Poole CD; Smith J, Davies JS. Cost-effectiveness and budget impact of Empirical vitamin D therapy on unintentional falls in older adults in the UK. (2015). BMJ Open. url: http://dx.doi.org/10.1136/bmjopen-2015-007910

4. Hiligsmann M, Sedrine WB, Bruyère O, et al. Cost-effectiveness of vitamin D and calcium supplementation in the treatment of elderly women and men with osteoporosis. (2015) European Journal of Public Health 25(1): 20–25. url: https://doi.org/10.1093/eurpub/cku119

5. Niedermaier T, Gredner T, Kuznia S. et al. Vitamin D supplementation to the older adult population in Germany has the cost-saving potential of preventing almost 30,000 cancer deaths per year. (2021). Mol Oncol. Online ahead of print. url: https://febs.onlinelibrary.wiley.com/doi/10.1002/1878-0261.12924

6. Central Statistics Office. IRISH LIFE TABLES NO. 17 2015-2017. url: https://www.cso.ie/en/releasesandpublications/er/ilt/irishlifetablesno172015-2017/

7. Central Statistics Office. Irish population statistics for 2016 by age/gender. Accessed on 09 Feb 2021. url: https://data.cso.ie/

8. Health Information and Quality Authority. Guidelines for the Economic Evaluation of Health Technologies in Ireland 2020. url: https://www.hiqa.ie/sites/default/files/2020-09/HTA-Economic-Guidelines-2020.pdf

9. Keum N, Lee DH, Greenwood DC, et al. Vitamin D supplementation and total cancer incidence and mortality: a meta-analysis of randomized controlled trials. (2019). Ann Oncol; 30(5): 733–743.

10. Yao P, Bennett D, Mafham M, et al. Vitamin D and Calcium for the Prevention of Fracture: A Systematic Review and Meta-analysis. (2019). JAMA Netw Open; 2(12): e1917789. url: https://jamanetwork.com/journals/jamanetworkopen/fullarticle/2757873

11. Kahwati LC, Palmieri Weber R, et al. Vitamin D, calcium or combined supplementation for the primary prevention of fractures in community-dwelling adults. Evidence report and systematic review for the US Preventative Services Task Force. (2018). JAMA; 319:1600–12.

12. Bischoff-Ferrari HA, Willett WC, Wong JB, et al. Prevention of nonvertebral fractures with oral vitamin D and dose dependency: a meta-analysis of randomized controlled trials. (2009). Arch Intern Med; 169(6):551–61.

13. National Office of Clinical Audit. IRISH HIP FRACTURE DATABASE NATIONAL REPORT 2019 - Stay safe and active at home. url: http://s3-eu-west-1.amazonaws.com/noca-uploads/general/Irish_Hip_Fracture_Database_National_Report_2019_10.11.2020.pdf

14. Kelly MA, McGowan B, McKenna MJ, et al. Emerging trends in hospitalisation for fragility fractures in Ireland. (2018). Ir Med J; 187(3): 601–8. url https://link.springer.com/article/10.1007/s11845-018-1743-z

15. Imai N, Endo N, Hoshino T, et al. Mortality after hip fracture with vertebral compression fracture is poor. (2016) J Bone Miner Metab. 34(1): 51–4. doi: 10.1007/s00774-014-0640-4. url: https://pubmed.ncbi.nlm.nih.gov/25501699/

16. Szende A, Janssen B, Cabases J (eds). In: Self-Reported Population Health: An International Perspective based on EQ-5D. (2014). Springer Open. (Table 3.6), pp 30. url: https://eq-5dpublications.euroqol.org/download?id=0_54006&fileId=54415

17. CANCER IN IRELAND 1994-2018 WITH ESTIMATES FOR 2018-2020: ANNUAL REPORT OF THE NATIONAL CANCER REGISTRY. 2020 Annual Report. url: https://www.ncri.ie/sites/ncri/files/pubs/NCRI_Annual%20Report_2020_01122020.pdf

18. Guest JF, Ruiz FJ, Greener MJ, et al. Palliative care treatment patterns and associated costs of healthcare resource use for specific advanced cancer patients in the UK. (2005). Eur. J. Cancer Care; 15(1):65–73. url https://onlinelibrary.wiley.com/doi/abs/10.1111/j.1365-2354.2005.00623.x

19. Primary Care Reimbursement Service (PCRS) Annual Report 2016. url: http://www.hse.ie/eng/staff/PCRS/PCRS_Publications/PCRS-Annual-Report-20161.pdf

20. National Institute of Clinical Care Excellence (NICE), Costing statement: Vitamin D: increasing supplement use among at-risk groups (PH56) (2014). url: https://www.nice.org.uk/guidance/ph56/resources/costing-statement-pdf-69288013

21. Primary Care Reimbursement Service (PCRS) database. url: https://www.sspcrs.ie/druglist/pub

22. CSO Irish health inflator from the Iris Central Statistics Office (CSO). url: https://data.cso.ie/

23. OECD. Purchasing Power Parities for GDP and related indicators. From stats. OECD.org. Accessed on 11 Feb 2021. url: https://stats.oecd.org/Index.aspx?DataSetCode=PPPGDP

24. Martineau AR, Jolliffe DA, Hooper RL, et al. Vitamin D supplementation to prevent acute respiratory tract infections: systematic review and meta-analysis of individual participant data. (2017) BMJ 356: i6583. url: https://doi.org/10.1136/bmj.i6583

